# Detection of asymptomatic SARS-CoV-2 infections among healthcare workers: results from a large-scale screening program based on rapid serological testing

**DOI:** 10.1101/2020.07.30.20149567

**Authors:** Francesca Maria Carozzi, Maria Grazia Cusi, Mauro Pistello, Luisa Galli, Alessandro Bartoloni, Gabriele Anichini, Chiara Azzari, Michele Emdin, Claudia Gandolfo, Fabrizio Maggi, Elisabetta Mantengoli, Maria Moriondo, Giovanna Moscato, Irene Paganini, Claudio Passino, Francesco Profili, Fabio Voller, Marco Zappa, Filippo Quattrone, Gian Maria Rossolini, Paolo Francesconi, SARS-CoV-2 serosurvey Tuscan working group

## Abstract

**Objective:** To evaluate the performance of two available rapid immunological tests for identification of severe acute respiratory syndrome Coronavirus 2 (SARS-CoV-2) antibodies and their subsequent application to a regional screening of health care workers (HCW) in Tuscany (Italy).

**Design:** measures of accuracy and HCW serological surveillance

**Setting:** 6 major health facilities in Tuscany, Italy.

**Participants:** 17,098 HCW of the Tuscany Region. Measures of accuracy were estimated to assess sensitivity in 176 hospitalized Covid-19 clinical subjects at least 14 days after a diagnostic PCR-positive assay result. Specificity was assessed in 295 sera biobanked in the pre-Covid-19 era in winter or summer 2013-14

**Main outcome measures:** Sensitivity and specificity, and 95% confidence intervals, were measured using two serological tests, named T-1 and T-2. Positive and Negative predictive values were estimated at different levels of prevalence. HCW of the health centers were tested using the serological tests, with a follow-up nasopharyngeal PCR-test swab in positive tested cases.

**Results:** Sensitivity was estimated as 99% (95%CI: 95%-100%) and 97% (95% CI: 90%-100%), whereas specificity was the 95% and 92%, for Test T-1 and T-2 respectively. In the historical samples IgM cross-reactions were detected in sera collected during the winter period, probably linked to other human coronaviruses. Out of the 17,098 tested, 3.1% have shown the presence of SARS-CoV-2 IgG antibodies, among them 6.8% were positive at PCR follow-up test on nasopharyngeal swabs.

**Conclusion:** Based on the low prevalence estimate observed in this survey, the use of serological test as a *stand-alone* test is not justified to assess the individual immunity status. Serological tests showed good performance and might be useful in an integrated surveillance, for identification of infected subjects and their contacts as required by the policy of *contact tracing*, with the aim to reduce the risk of dissemination, especially in health service facilities.

## Introduction

In Italy the first autochthonous cases of coronavirus disease 2019 (COVID-19) were identified in two geographical areas of Northern Italy in mid-February 2020 and thereafter the disease spread to other regions with a north-south gradient. Although the different severity in severe acute respiratory syndrome coronavirus 2 (SARS-CoV-2) propagation, the containment measures were extended to the whole country since 11th of March 2020. The lockdown policy resulted in curbing the epidemic curve and was concluded on 4th of May, when the early post-lockdown phase started (1).

Diagnostic tests for severe infections are currently divided into two main categories: those that detect the presence of SARS-CoV-2 and are primarily used to diagnose an active infection, and those that detect the presence of antibodies against SARS-CoV-2. World Health Organization (WHO) recommended to firstly test patients showing SARS-related signs and symptoms and an history of travelling to epidemic areas or contact with known or suspected SARS-CoV-2 infected subject (2), and only later to extend the analysis to asymptomatic or paucisymptomatic subjects. As currently recommended by the WHO, routine confirmation of COVID-19 cases in suspected subjects is based on the detection of unique viral RNA sequences, by nucleic acid amplification tests (NAAT), such as RT-PCR, following by confirmation using nucleic acid sequencing when necessary or feasible. Preliminary reports on sensitivity ranged from 27% to 98%, while specificity is claimed to be very high (3). The laboratory process is quite complex and results are generally available within 24 hours, although testing large numbers of subjects is posing a great strain on facilities.

Most cases tested in the course of the first phase of the epidemic have been acutely ill and highly symptomatic, while asymptomatic or mildly symptomatic individuals have more rarely been tested. Limited test availability, especially at the beginning of Italian epidemic, and preferential testing of symptomatic patients, likely led to underestimation of the infection burden and overestimation of fatality rates (4, 5). The PCR-test positivity in nasopharyngeal swabs usually persists for a median of 2-3 weeks in infected subjects but it may remain positive for a longer period (median 30 days) and even after the symptoms have disappeared (6).

The use of rapid antibody tests has been considered for large scale screening of at-risk populations in order to assess past exposure to SARS-CoV-2 and identify asymptomatic viral carriers (7). A testing strategy capable of reliably detecting subjects who have been (knowingly or unknowingly) infected and successfully recovered from the infection would allow to obtain a more accurate estimate of the prevalence of the disease.

At the present, the antibody kinetics against the SARS-CoV-2 is mostly unknown. Zhang et al (6) and Long et al (8) found viral antibodies in near all patients with COVID-19 infections, a marker of the transition from earlier to late period of infection. Timing at first antibody detection is dependent on the sensitivity of the method and the viral protein used as an antigen (6). The census of rapid serological tests for the detection of IgG and IgM available on the world market includes more than 100 different kits and is growing. The analytical performance of the individual commercially available diagnostic kits is still largely unknown. Identifying manufacturer and distributor of kits is not always possible and scientific publications, not commercial reports, assessing measures of performance are few. Recently a Cochrane Systematic Review (9) reported that sensitivity has mainly been evaluated in hospitalized patients, so it is unclear whether the tests are able to detect lower antibody levels likely associated with milder and asymptomatic COVID□19 disease. Therefore, before starting with epidemiological studies, assessment of analytical and clinical performances in independent studies are needed.

In Italy, the contribution of healthcare workers (HCW) in terms of COVID-19 cases and deaths has been relevant. A crucial step in limiting diffusion of infection is represented by detecting of infected subjects who remain asymptomatic and therefore may represent a significant source of cross-infection (10). This aspect is especially relevant in the healthcare settings, where infection can spread rapidly and involve both HCW and patients. In April 2020, the Regional Health Department of Tuscany Region promoted a large-scale serological surveillance of HCWs, aimed to identify asymptomatic subjects at risk of SARS-CoV-2 infection. In the present study, we report the analytical and clinical performances of the rapid serological tests used in this survey, the seroprevalence of SARS-CoV-2 antibodies in a large group of HCWs in Tuscany Region, and the identification of asymptomatic viral carriers among seropositive subjects.

## Methods

In Tuscany Region, according with the ordinance of the Health Care Department, rapid serological testing for anti-SARS-CoV-2 IgG and IgM antibodies was offered to HCWs during April 2020.

The participation in the survey was on a voluntary basis, and all subjects tested positive were invited to perform swab testing to detect the presence of SARS-CoV-2 via real-time reverse-transcription polymerase chain reaction (RT-PCR).

The present report describes the results of the survey carried out on HCWs of the four Tuscan University Hospitals (AOUS in Siena, AOUC in Florence, AOUP in Pisa, and AOUM, the Meyer Children’s University Hospital, in Florence) and of two additional healthcare Institutions, the Institute for prevention, research and oncological network (ISPRO) in Florence and the Fondazione Toscana Gabriele Monasterio (FTGM) in Pisa and Massa. A code was assigned to each participating Center as follows: Site-A, Site-B, Site-C, Site-D, Site-E and Site-F.

### Rapid serological tests (RST) for anti-SARS-CoV-2 IgG and IgM

Two rapid serological tests (RST) (Screen Test Covid-19 2019-nCOV IgG/IgM by Screen Italia S.r.l and COVID-19 IgG/IgM rapid test cassette by Zhejiang Orient Gene Biotech Co., Ltd) for the detection of anti-SARS-CoV-2 antibodies were used in the present study for large-scale screening and evaluated to assess sensitivity and specificity. The two immunological tests were provided by the Health Regional Department and they are subsequently referred as Test T-1 and Test T-2, respectively. Both tests were rapid lateral flow immunochromatographic assays for the qualitative detection of IgG and IgM antibodies directed to SARS-CoV-2 in human whole blood, serum or plasma. Test T-1 was performed using 20 μL of whole blood or 10 μL of serum/plasma dispensed together with two drops of buffer in the same single well located in the cassette. Test T-2 was performed using 10 μL of whole blood or 5 μL of serum/plasma, placing them in the proximal well of the cassette and adding two drops of the buffer to the buffer well. An internal quality control was present in both tests.

#### Results were interpreted as follows

Negative results: presence of the expected control line with no lines at the IgG and IgM positions.

IgG positive: presence of the expected control line and of a line at the IgG position only.

IgM positive: presence of the expected control line and of a line at the IgM position only.

IgG and IgM positive: presence of the expected control line and of two lines at the IgG and IgM positions, respectively.

The IgG or IgM readings were considered doubtful if a shade, not classifiable as a clear line, appeared at the IgG or/and IgM positions.

The test was considered invalid in the absence of the expected control line.

The RST were performed in six laboratory departments of the participating institutions. Before starting, reading procedures of the rapid tests were defined and shared.

### Sensitivity and Specificity Evaluation

Sensitivity of the RST in detecting anti-SARS-CoV-2 antibodies was evaluated considering sera from virologically-confirmed COVID-19 symptomatic patients at advanced stages of the disease as true positives. In particular, 109 sera collected at least 14 days after the beginning of symptoms (68 from Site-C and 41 from Site-B were considered for validation of test A); the same 41 sera from Site-B used for validation of test T-1, were added to 26 sera, collected at least 10 days after the beginning of symptoms at Site-A for the validation of test T-2.

Specificity was evaluated at Site-F, using 295 anonymous sera collected in the pre-COVID era (2013-2014) and considered as true negatives for the presence of anti-SARS-Cov-2 antibodies. The 295 sera were collected from 145 women and 150 men aged 50-70, equally distributed among gender; among the 295 sera, 200 (100 from men and 100 from women) were collected during winter months (from November to February) and 95 (50 from men and 45 from women) during summer months (July and August).Test T-1 and test T-2 were performed simultaneously and the results were interpreted by the same operator.

### Molecular testing for SARS-CoV-2 in nasopharyngeal swabs

A nasopharyngeal PCR-test swab was collected using eSwab® device (Copan Italy) from HCWs who received a positive serological test result within 3 days from serological testing. Viral RNA was extracted from nasopharyngeal swabs using an automated system (NIMBUS,Seegene). The detection of SARS-Cov-2 RNA was performed by RT-real time PCR (Allplex™ 2019-nCoV Assay, Seegene) amplifying three viral genes (E, RdRp and N) and an internal control (IC). The amplification occurred if Ct was inferior to 40 cycles.

#### Results were interpreted as follows

Viral RNA was considered as not detected if only the IC was amplified, as detected if at least one of the viral targets was amplified with a Ct≤35, and as detected with low viral load if only one of the viral targets was amplified with a CT>35.

### Study design for large scale screening of healthcare workers and detection of asymptomatic infections

The Tuscany Region screening was directed to all healthcare workers in public structures. The study was approved by the local Ethic Committees and offered voluntarily to all personnel who underwent a rapid serological test at their institution.

Test T-1 was adopted at Site-B while all other institutions used immunological Test T-2. All subjects resulted positive to the serological test were then invited to perform a PCR-test swab within 3 days, except Site-A adopting a different assessment protocol (not included in this paper). In this analysis, subjects were not classified for risk of having contracted COVID-19 due to their professional function nor a detailed information of previous symptoms linked to COVID-19 infection was collected.

Accuracy measure estimates and their 95% confidence intervals were calculated using the binomial distribution with the Stata 12.0 software. A glossary of statistical measures and definitions of the measures are presented in BOX 1. According to MHRA (11), we estimated specificity considering only positive or dubious IgG results, regardless IgM status.

**BOX 1.**
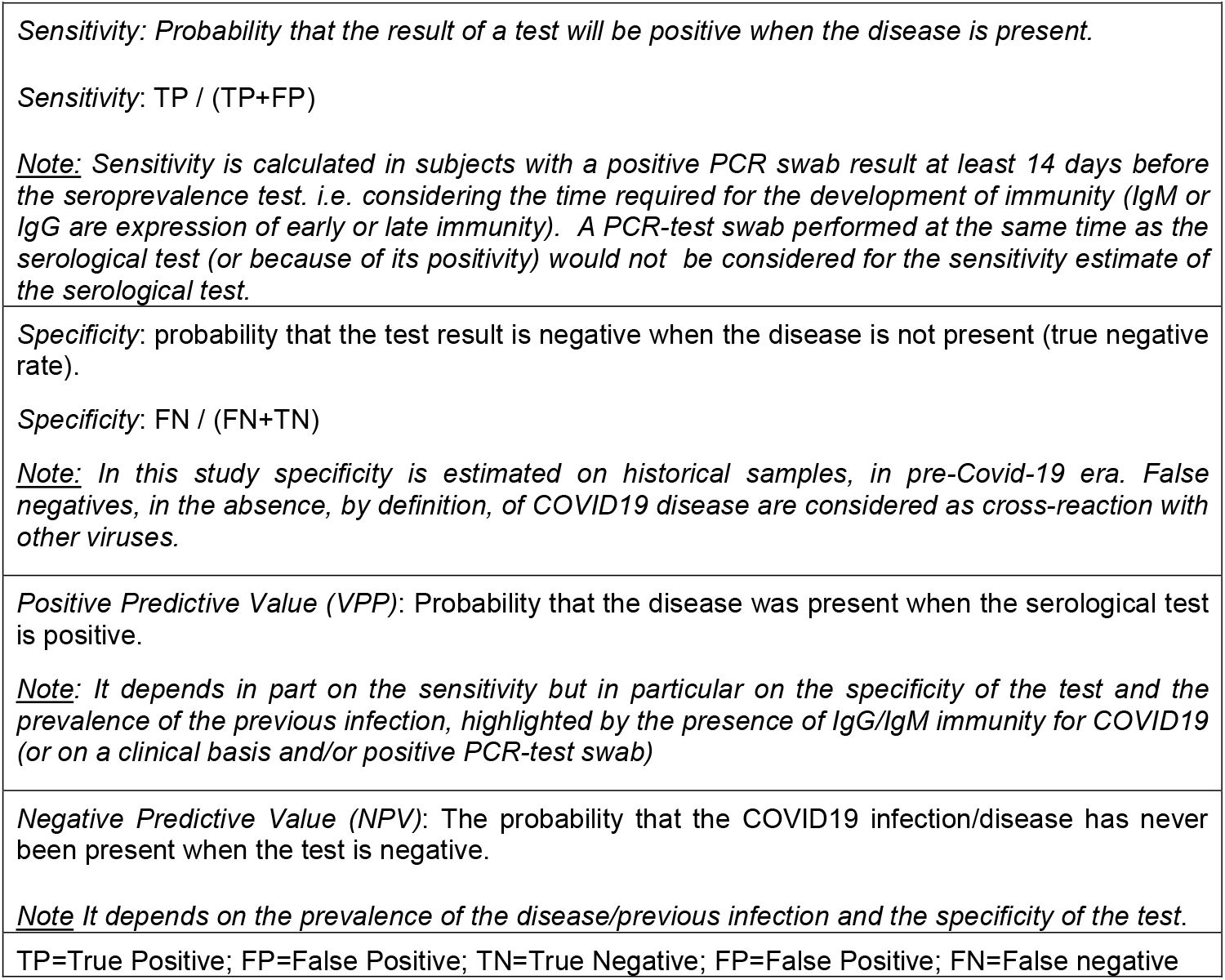
Glossary.

|  |
| --- |
| **BOX 1 – Glossary**<br>*Sensitivity: Probability that the result of a test will be positive when the disease is present.*<br>*Sensitivity: $TP / (TP+FP)$*<br>*<u>Note:</u> Sensitivity is calculated in subjects with a positive PCR swab result at least 14 days before the seroprevalence test. i.e. considering the time required for the development of immunity (IgM or IgG are expression of early or late immunity). A PCR-test swab performed at the same time as the serological test (or because of its positivity) would not be considered for the sensitivity estimate of the serological test.* |
| *Specificity: probability that the test result is negative when the disease is not present (true negative rate).*<br>*Specificity: $FN / (FN+TN)$*<br>*<u>Note:</u> In this study specificity is estimated on historical samples, in pre-Covid-19 era. False negatives, in the absence, by definition, of COVID19 disease are considered as cross-reaction with other viruses.* |
| *Positive Predictive Value (VPP): Probability that the disease was present when the serological test is positive.*<br>*<u>Note:</u> It depends in part on the sensitivity but in particular on the specificity of the test and the prevalence of the previous infection, highlighted by the presence of IgG/IgM immunity for COVID19 (or on a clinical basis and/or positive PCR-test swab)* |
| *Negative Predictive Value (NPV): The probability that the COVID19 infection/disease has never been present when the test is negative.*<br>*<u>Note</u> It depends on the prevalence of the disease/previous infection and the specificity of the test.* |
| TP=True Positive; FP=False Positive; TN=True Negative; FP=False Positive; FN=False negative |

## Results

### Evaluation of the performance of RST used in this study

Sensitivity of the two RST for the detection of anti-SARS-CoV-2 antibodies was evaluated using serum samples from virologically confirmed COVID-19 patients reporting relevant signs and symptoms since 10 - 14 days. Overall, the sensitivity of the Test T-1 and T-2, calculated on 67 and 109 serum specimens, respectively, and considering positivity for any Ig class, was found to be 99% (95%CI: 95-100%) and 97% (95% CI: 90-100%), respectively. Considering the different Ig classes, sensitivity was 82% (95%CI: 73-88) and 94% (95%CI: 85-98) for IgG and 72% (95%CI: 63-81) and 82% (95%CI: 71-90) for IgM, respectively (Table 1).

**Table 1:**
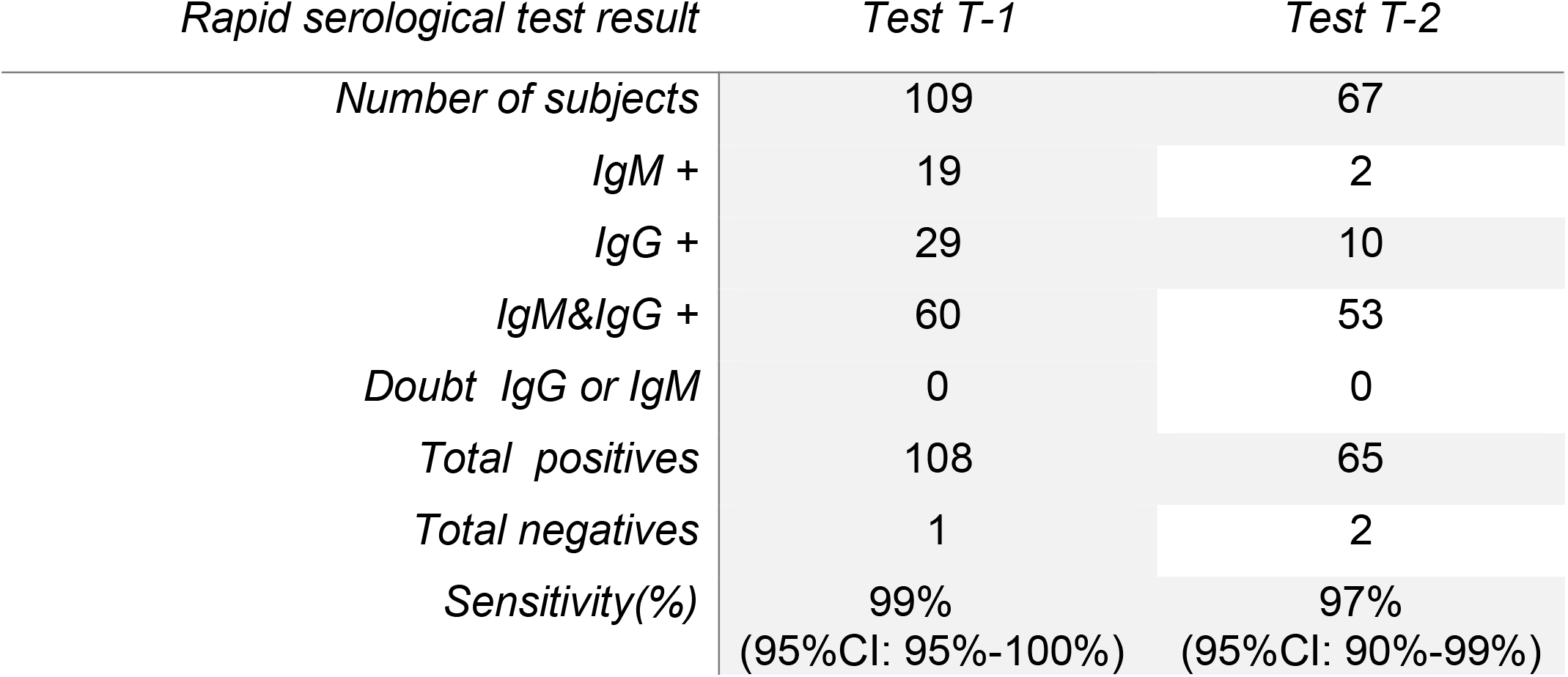
Sensitivity of serological rapid tests. Test performed in sera from clinical hospitalized COVID-19 cases after 14 days since a positive PCR-test swab

Specificity of the two RST was evaluated using 295 archival serum samples collected in the pre-COVID era. Overall, the specificity of the Test T-1 and Test T-2 for any Ig was 83% and 88% respectively. Considering only the IgG class, the specificity was 95% (95% CI: 92%-97%) for Test T-1 and 92% (95% CI: 89%-95%) for test T-2. Test T-1 showed a higher frequency of doubtful results for IgM in comparison with Test T-2 [8% (25/295) vs 1.7% (5/295), p-value<0.0001, but a lower proportion of cross-reactions for IgG [4.7% (14/295) for test T-1 vs 7.8% (23/295) for test T-2, p-value=0.0471]. Figure 1 shows the discordant results by test T-1 and T-2. No cross-reaction was detected in the same sample for test A or B, except in two cases.

**Figure 1:**
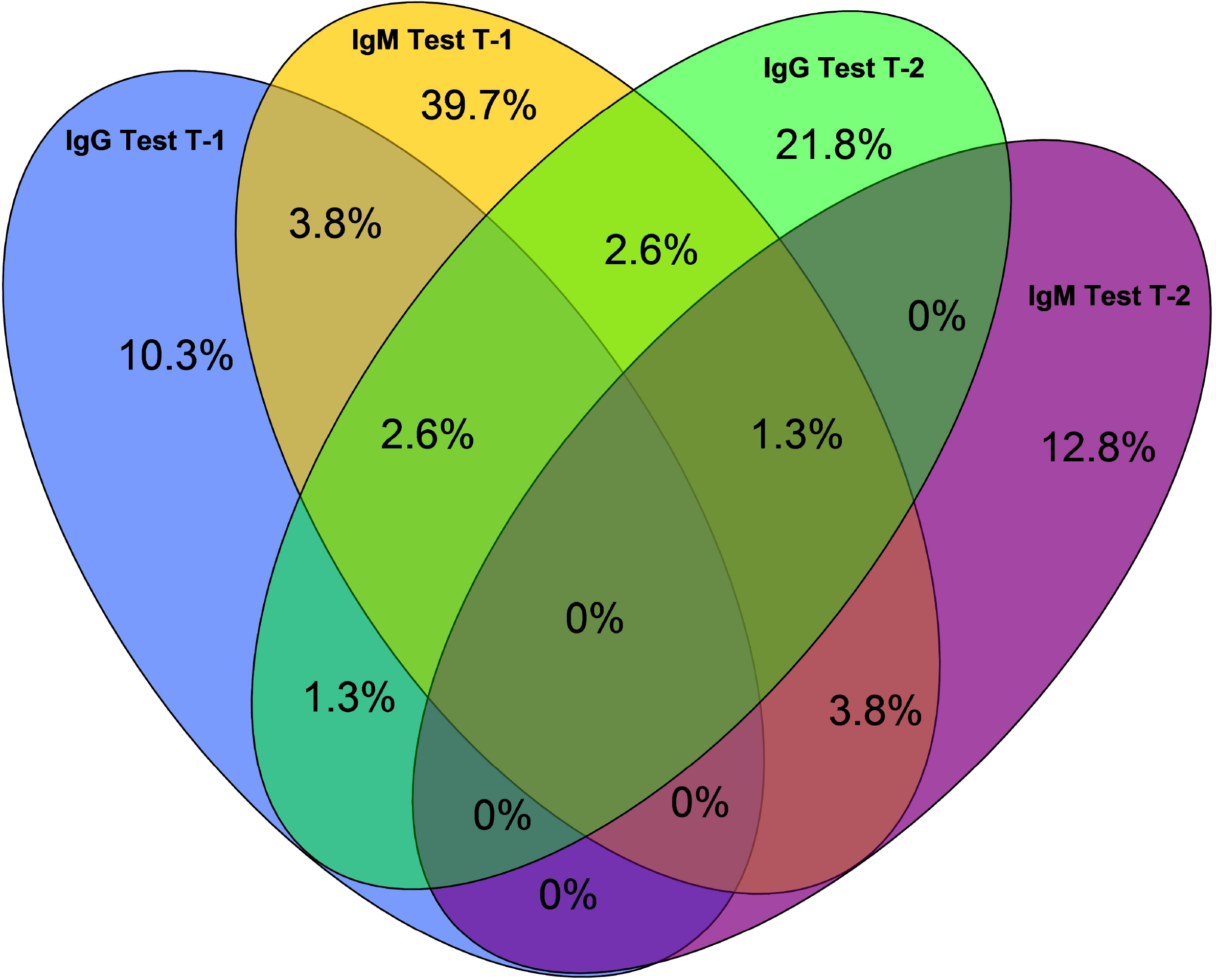
Disagreement in rapid serological test results by Test T-1 and T-2. Based on all the 295 pre-COVI19 historical sera - Proportions were calculated from all samples that tested positive. Doubt results were considered positive.

In Table 2 specificity measure is showed by seasonal period of sample’s collection (200 samples in winter and 95 in summer). In total the specificity of IgG positives or dubious was 95% and 92%, for Test T-1 and Test T-2, respectively, with similar values by season. On the contrary, specificity for IgM was significantly lower in winter for both tests (p=0.0073 and p=0.0397), showing an higher proportion of cross reactions possibly due to seasonal viral infections.

**Table 2:** Specificity of serological rapid tests. Historical bio-banked sera (2013-14), by season of collection. False Positive tests are cross-reactions

| *Rapid serological test result* | ***Test T-1*** |  |  | ***Test T-2*** |  |  |
| --- | --- | --- | --- | --- | --- | --- |
|  | *Total* | *Winter* | *Summer* | *Total* | *Winter* | *Summer* |
| *Number of subjects* | 295 | 200 | 95 | 295 | 200 | 95 |
| *IgM + or doubt* | 37 | 32 | 5 | 13 | 12 | 1 |
| *IgG + or doubt* | 9 | 6 | 3 | 22 | 15 | 7 |
| *IgM & IgG + or doubt* | 5 | 4 | 1 | 1 | 1 | 0 |
| *IgG negatives* | 281 | 190 | 91 | 272 | 184 | 87 |
| *Specificity for any Ig* | 83% | 79% | 91% | 88% | 86% | 92% |
| *IgG Specificity (%)* | 95% | 95% | 96% | 92% | 92% | 93% |

Table 3 shows the positive predictive value (PPV) and negative predictive value (NPV) calculated at different prevalence for both tests (Table 3a and 3b for Test-T1 and Test-T2, respectively), given the estimated values of sensitivity and specificity. NPV was high (99.9%) for both tests, regardless the prevalence. At low level of prevalence (2%), PPVs was estimated at 30% and 20% for test T-1 and Test T-2, respectively, while with higher prevalence (10%) PPVs increased at 68% and 57%, respectively.

**Table 3a:** Sensitivity, specificity and positive and negative predictive values (PPV, NPV) of serological Test T-1 at different level of actual prevalence.

|  |  |  |  |  |  |  |
| --- | --- | --- | --- | --- | --- | --- |
| **Sensitivity** | 99% (95% CI: 95%-100%) |  |  |  |  |  |
| **Specificity** | 95% (95% CI: 92%-97%) |  |  |  |  |  |
| **ACTUAL PREVALENCE** | **1%** | **2%** | **3%** | **5%** | **10%** | **20%** |
| **PPV** | 17% | 30% | 39% | 52% | 68% | 83% |
| **NPV** | 100% | 100% | 100% | 100% | 99% | 98% |

**Table 3b:** Sensitivity, specificity and positive and negative predictive values (PPV, NPV) of serological Test T-2 at different level of actual prevalence.

|  |  |  |  |  |  |  |
| --- | --- | --- | --- | --- | --- | --- |
| **Sensitivity** | 97% (95% CI: 90%-100%) |  |  |  |  |  |
| **Specificity** | 92% (95% CI: 89%-95%) |  |  |  |  |  |
| **ACTUAL PREVALENCE** | **1%** | **2%** | **3%** | **5%** | **10%** | **20%** |
| **PPV** | 11% | 20% | 28% | 39% | 57% | 75% |
| **NPV** | 100% | 100% | 100% | 100% | 99% | 98% |

### Large-scale screening of Tuscan HCWs for SARS-CoV-2 serological reactivity and asymptomatic SARS-CoV-2 infections

A total of 17,098 HCWs from the six healthcare facilities participating in the study underwent screening with RST during April 2020. Table 4 shows the results of the serological survey by facility. Overall, the rate of seropositivity (considering IgM and/or IgG positive or doubtful results) was 3.4%. Overall, the rate of doubtful results for seropositivity was 0.8%.

**Table 4:**
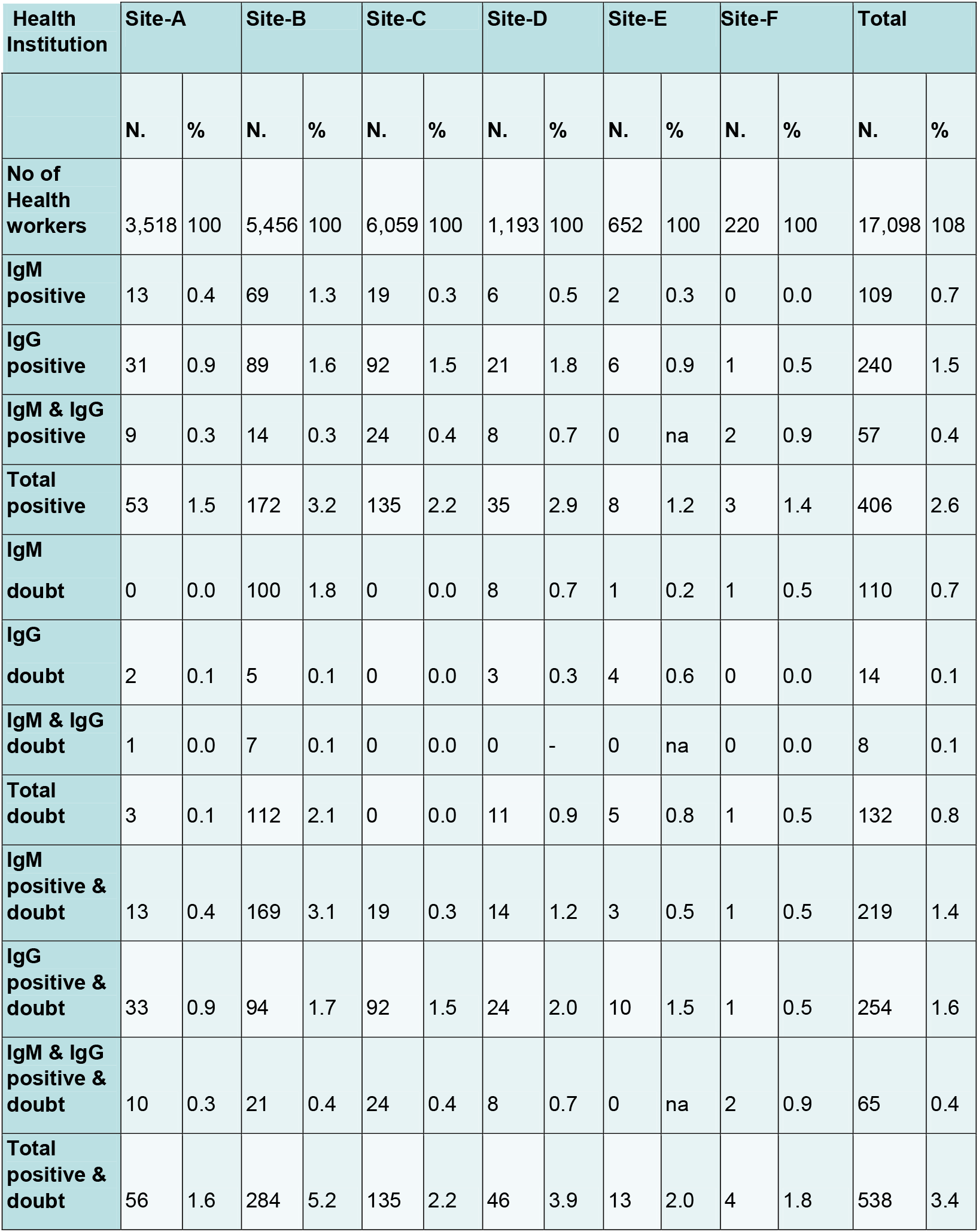
Results of serological tests in health care workers of the Tuscany Region. by institution.

The proportion of seropositivity ranged from 1.6 to 5.2% by health institution, (p<0.001). The highest seropositivity rate and the highest rate of doubtful results were reported at Site-B (2.1%), where test A was used. Excluding doubtful results, most of the positive results were due to IgG (73.1%), as compared to IgM (40.9%).

Table 5 shows the number and proportion of HCWs positive to PCR-test swabs among those positives to serological rapid test. Excluding Site-A, where a different recall protocol was adopted, most of the HCWs that resulted positive to RST underwent molecular testing for SARS-CoV-2 RNA in a nasopharyngeal swab within three days since serological testing. Overall, for 33 of the seropositive subjects detected at sites B-F (6.8%) viral RNA was detected. The rate of viral positivity among seropositive subjects ranged from 0% (Site-F) to 15.4% (Site-E). The results by sites were not statistically different (chi-square = 4.18; p=0.38)

**Table 5:** Results of PCR-test swab in health care workers of the Tuscany Region with positive serological test (N=403).

|  | Site-B<br>(N=284) |  | Site-C<br>(N=135) |  | Site-D<br>(N=46) |  | Site-E<br>(N=13) |  | Site-F<br>(N=4) |  | Total<br>(N=403) |  |
| --- | --- | --- | --- | --- | --- | --- | --- | --- | --- | --- | --- | --- |
|  | PCR-test<br>positive | % | PCR-test<br>positive | % | PCR-test<br>positive | % | PCR-test<br>positive | % | PCR-test<br>positive | % | PCR-test<br>positive | % |
| IgM pos | 1 | 1.4 | 0 | 0.0 | 0 | 0.0 | 0 | 0 | 0 | 0.0 | 1 | 1.0 |
| IgG pos | 14 | 15.7 | 1 | 1.1 | 2 | 9.5 | 2 | 33.3 | 0 | 0.0 | 19 | 9.1 |
| IgM & IgG<br>pos | 4 | 28.6 | 5 | 20.8 | 2 | 25.0 | 0 | 0.0 | 0 | 0.0 | 11 | 22.9 |
| Total<br>positive | 19 | 11.0 | 6 | 4.4 | 4 | 11.4 | 2 | 25.0 | 0 | 0.0 | 31 | 8.8 |
| IgM doubt | 0 | 0.0 | 0 | 0.0 | 0 | 0.0 | 0 | 0.0 | 0 | 0.0 | 0 | 0.0 |
| IgG doubt | 0 | 0.0 | 0 | 0.0 | 1 | 33.3 | 0 | 0.0 | 0 | 0.0 | 1 | 8.3 |
| IgM & IgG<br>doubt | 1 | 14.3 | 0 | 0.0 | 0 | na | 0 | 0.0 | 0 | 0.0 | 1 | 14. |
| Totale doubt | 1 | 0.9 | 0 | 0.0 | 1 | 9.1 | 0 | 0.0 | 0 | 0.0 | 2 | 1.6 |
| IgM pos &<br>doubt | 1 | 0.6 | 0 | 0.0 | 0 | 0.0 | 0 | 0.0 | 0 | 0.0 | 1 | 0.5 |
| IgG pos &<br>doubt | 14 | 14.9 | 1 | 1.1 | 3 | 12.5 | 0 | 0.0 | 0 | 0.0 | 20 | 9.0 |
| IgM & IgG<br>pos and<br>doubt | 5 | 23.8 | 5 | 20.8 | 2 | 25.0 | 0 | 0.0 | 0 | 0.0 | 12 | 21.8 |
| Total<br>positive and<br>doubt | 20 | 7.0 | 6 | 4.4 | 5 | 10.9 | 2 | 15.4 | 0 | 0.0 | 33 | 6.8 |

Note: Site-A not included because of the different assessment protocol.

## Discussion

Several diagnostic strategies are available to identify an ongoing infection, to rule out an infection or to test for past infection by SARS-CoV-2 and immune response to the virus.

Serology testing to detect the presence of anti-SARS-CoV-2 antibodies aims firstly to identify previous SARS-CoV-2 exposure and assess the prevalence of subjects that have experienced SARS-CoV-2 infection and developed immunity. Moreover, serology testing can also help to detect an ongoing infection because the presence of specific antibodies may be detected either in symptomatic or asymptomatic subjects when viral markers are still present. The sensitivity of antibody testing is too low in the first week since symptom onset; therefore they cannot have a primary role for the diagnosis of COVID-19, but they may still maintain a role for integrating other tests when RT-PCR cannot be performed (6).

At present, the antibody kinetics against SARS-CoV-2 antigens is not completely known, but several studies have shown that specific IgM and IgG antibodies can be detected as early as the 4th day after symptoms onset (12, 13). However, the time at antibody detection is dependent on the sensitivity of the method. Tan et al. (14) reported that the anti-nucleocapsid-protein IgM become positive on day 7 and the positivity rate peaked on day 28, while the IgG compared on day 10 and reach their peak at day 49 after disease onset.

The use of serological antibody testing has been promoted in many Italian areas as a tool for surveillance. Considering that HCWs are both more exposed to the COVID-19 infection and a source of infection for patients, the Tuscany regional government decided for the implementation of a mass seroprevalence survey of these job categories. A volunteer mass screening by means of immunological tests was first offered to workers of the University Hospitals of the Tuscany Region and afterwards to all other regional health institutions. To identify asymptomatic infections, all HCWs who resulted IgG or IgM positive or doubtful were as soon as possible and in any case within three days tested for SARS-CoV-2 RNA and, if positive, immediately isolated.

Immunological tests were provided by the Health Regional Department in order to offer surveillance to the HCWs. Despite serological screening based on CLIA or ELISA were already available, rapid serological tests, exploiting lateral immunochromatographic flow, were considered more suitable for mass screening. Before the large-scale survey, an evaluation of the performances of the two available kits was carried out. Sensitivity and specificity of both tests (A and B) were shown to be acceptable on the basis of the MHRA document (11), which prescribed at least 98% for both measures within the 95% CI of each measure. This level of performance was also considered adequate for the approval of rapid tests at 95% of sensitivity and specificity by FDA.

In a recent study analyzing data from the Chinese (12) COVID-19 subjects, acute antibody responses to SARS-CoV-2 were shown within 19 days after symptom onset, with a 100% of patients tested positive for antiviral IgG. Seroconversion occurred sequentially. The study confirmed that asymptomatic COVID-19 cases represent a relevant problem. In our study both Ig classes (IgG and IgM) showed high sensitivity in subjects with clinical COVID-19, however sensitivity of test T-1 and test T-2 resulting from asymptomatic infections, could not be evaluated, since the true sensitivity of the test appeared to be overestimated.

Testing sera bio-banked in the pre-COVID19 era, the positive results could not be attributable to SARS-CoV-2. A lower specificity in samples collected during the winter period was observed when compared with the ones collected in summer. The seasonal lower specificity was mainly due to IgM positivity, which likely reflected cross-reactivity with other human coronaviruses or other viruses circulating during winter season.

Screening of more than 17,000 HCWs by serological test for SARS CoV-2 showed a prevalence of 3.4%, in Tuscany. Variability of exposure among centers might suggest different levels of risk of the HCWs, who were representative of all the different activities carried out in health facilities. The number of HCWs resulting positive to serological tests was, on average, lower than expected, considering the high risk of job categories but probably, availability and correct use of Personal protective equipment (PPE) played an important role. Among them, the number of positive to PCR-test swab was rare (2,4 % of all screened subjects). Serological tests could contribute to identify a small number of asymptomatic HCWs who can spread the infection and determine a potential nosocomial outbreak.

A negative IgG result is considered an indicator for the absence of immunity. On the basis of the serological tests the possibility of an actual infective status, before the seroconversion, is possible. The probability of a false negative result depends on the epidemic curve, i.e. the risk of identification of a subject classified as non-infected whereas in latent phase is related to the chance of contagion. In the presence of a low level of virus circulation among the target population, the large majority are expected to be true negative results. The opportunity of test repetition might be considered after one week or a month, resulting in a potentially higher sensitivity of the test (15).

How to deal with a positive serological test is still an open question (16). There are many uncertainties about the immunity, the test accuracy and the possible consequences caused by a positive misclassification. However, during the epidemic spread, an apparent benefit/harm ratio had to be taken into account in the grade of certainty. In the presence of a low prevalence, even with a good test performance, the estimate of the true prevalence is critical. A simple rule suggests the estimate of true prevalence is near zero when the actual prevalence is lower than (1-minus) specificity. Based on the prevalence estimate in this survey, the use of serological test as a *stand-alone* test is then not justified to assess the individual immunity status. Serological tests which have shown good performance must be used in the context of an integrated surveillance including an adequate contact tracing policy and general infection and prevention control measures for respiratory viruses. First, a PCR-test swab is a way to confirm the COVID-19 positivity, supporting for an early pauci- or asymptomatic infection. Positivity at PCR-test is quite rare in this large series of HCWs, and the large majority of those resulted positive to the serological test was found to be negative at the virological assessment, i.e. they might be false positives or show a mature immunity in regard to SARS-CoV-2.

The ultimate result of this large serosurvey, is an integrated strategy for an optimized use of the serological testing, which should be validated (Table 6). Integration of policies is important for the early identification of pauci-or asymptomatic cases. If the test is used with the purpose to assess the personal immunity, false reassurance *per se* and others should be reduced as much as possible. Testing might be used as a *proxy* to suspect an ongoing or previous asymptomatic infection (especially in the presence of IgG) in mass screening programs, but it needs careful evaluation and follow up before considering it a warranty of individual immunity.

**Table 6:** Integrated strategy for serologic surveillance of high-risk populations.

| Rapid test Result | Rational | Action | Sub-action | Indicator(s) | Note |
| --- | --- | --- | --- | --- | --- |
| **Negative IgG & IgM** | Low probability of actual infection, in the absence of reported contacts or COVID19 symptoms.<br>Possible status of early asymptomatic infection (False Negative) | Repeat rapid test: timing is to relate to epidemic curve monitoring<br>With a new symptom or contact, a PCR-Test swab. | If negative at repeated rapid serological test continue testing at scheduled timing (relate to epidemic curve ) until surveillance continue.<br>Suggested GP counseling for symptoms and contact Tracing, if needed. | Number and proportion of IgG or IgM seroconversion at repeated rapid Test.<br>Frequency of intercurrent symptoms or contacts | A confirmed. negative Rapid Test is a likely indicator of the absence of a previous infection.<br>False Negative occurrence is unknown.<br>New. early Infections (also asymptomatic) must be identified early using repeated rapid test testing, clinical symptoms or contact tracing. PCR-test is the gold standard to assess infectivity.<br>. |
| **Positive IgG&/or IgM or dubious** | It's an indicator of a possible previous COVID19 infection.<br>A False Positive subject if assumed as immune to COVID19 is dangerous per se and others (false reassurance) | PCR-test swab to all serological positives. | If immediate PCR-test swab is positive, there is strong probability of actual disseminating infection. Surveillance and contact tracing is needed.<br>IF immediate PCR swab is negative, rapid serological test control after 7-14 days. Optional PCR-test swab repetition. | Number and proportion of IgG/IgM or dubious positive subjects confirmed as PCR-test positive.<br>Number of IgG/IgM positive subjects confirmed as PCR-test negative after 2 rapid serological test positive. | At the end of the surveillance complete protocol the subject is could be considered as probably immune.<br>Limited knowledge exists about the neutralizing action of antibodies, the possible limited duration of the immunity and possibilities of recurrences of COVID-19. Policy will be updated according with new knowledge. |

## Data Availability

All data referred into the manuscript are available.

